# VAERS data reveals no increased risk of neuroautoimmune adverse events from COVID-19 vaccines

**DOI:** 10.1101/2021.06.13.21258851

**Authors:** Chris von Csefalvay

## Abstract

Neuroautoimmune disorders, such as multiple sclerosis and Guillain-Barre syndrome, have been documented in relation to various vaccines in the past. This paper uses passive reporting information from the CDC/FDA’s VAERS system to analyse whether neuroautoimmune presentations are reported at a relatively higher or lower rate, vis-a-vis other adverse effects, for COVID-19 vaccines than for other vaccines. Through computing the reporting odds ratios for a range of symptoms and comparator vaccines, a clear indication in favour of the safety of COVID-19 vaccines emerges, with reports of neuroautoimmune adverse events in relation to other adverse events being over 70% less likely for COVID-19 than for comparator vaccines (*ROR* : 0.292, *p* < 0.0001). In comparison with other vaccines given as part of routine care in adulthood, COVID-19 vaccines have the lowest reporting odds ratio of neuroautoimmune adverse effects (median *ROR*: 0.246).

## 1. Introduction

The global drive to curb the spread of COVID-19 through a concerted vaccination effort is undoubtedly one of the most ambitious public health projects ever undertaken. At the time of writing, only a little over six months after the FDA granted EUAs to the first two of eventually three SARS-CoV-2 vaccines in early December, over 43% of the U.S. population has been fully vaccinated (i.e. two doses of a two-dose vaccine or a single dose of a one-dose vaccine). For comparison, during the severe 2017-18 flu season, only 37.1% of adults were vaccinated.[1]

To safeguard this unprecedented accomplishment, it is crucial to exercise close surveillance of potential adverse events (AEFIs). At least partially owing to the fact that the Moderna and Pfizer/BioNTech vaccines are first-in-man applications of mRNA vaccine technology at scale, a significant number of eligible individuals have refused to be vaccinated.[2–4] The fear of adverse effects, both short and long term, is undoubtedly a driving factor that fuels these apprehensions and eventually results in an overall reluctance to be vaccinated. Among mid- to long-term AEFIs, neurological AEFIs hold a particular place, as is evident from studies on reasons behind vaccine refusal.[5] The bitter past experience with Guillain-Barre syndrome (GBS) as a side effect of certain influenza vaccines, esp. the much publicised association between 1976-77 A/New Jersey/8/76 influenza vaccines and GBS, is still within living memory.[6] Similarly well-known is the association between the recombinant hepatitis B vaccine and multiple sclerosis.[7] In addition, the emergence of various theories about a purported (and disproven) causal mechanism between autism spectrum disorders and vaccinations that hypothesises an autoimmune pathogenetic process has created a persistent point of rhetoric that erroneously portrays vaccines as inevitably likely to cause neuroautoimmunity.[8] Given the often devastating economic, social, psychological and QoL impact of neuroautoimmune diseases,[9–11] some degree of apprehension appears natural.

It is beyond the scope of this paper to outline the range of potential pathophysiological mechanisms that may underlie autoimmunity, including neuroautoimmunity, following a vaccination (for such a review, see Wraith et al. (2003)[12]). Rather, we seek to identify evidence regarding the safety of COVID-19 vaccines approved in the United States (Pfizer/BioNTech, Moderna and Johnson & Johnson) with regard to neuroautoimmune AEFIs through an analysis of VAERS.

As with all studies leveraging VAERS, its limitations must be read in conjunction with results derived from VAERS data. Like all passive pharmacovigilance systems, it relies on reporting. Under the terms of the EUAs granted to each of the three vaccine manufacturers, COVID-19 vaccination providers must report any

- vaccine administration errors (regardless of consequence);
- serious AEFI (regardless of causal attribution), incl. any AEFI resulting in inpatient admission, long-term disability or death; and
- instances of multi-system inflammatory syndrome,

along with serious immunisation failures (cases of COVID-19 in vaccinated individuals that result in inpatient admission or death). This goes beyond to the typical reporting regime, wherein reporting is encouraged but not mandatory unless the patient was a minor at the time. At the same time, VAERS remains open to reports from individuals, allied healthcare workers and even unrelated third parties.[13–15] In other words, while there is a risk of unreported AEFIs, there may also be a degree of over-reporting (the same AEFI, for instance, may be reported by the physician, the nurse and the patient as well, without the knowledge of any of the other parties). Moreover, VAERS does not verify the accuracy or veracity of reports, nor does it require a causal attribution. This is notably illustrated by the number of accidents, drownings and congenital diagnoses reported to VAERS, none of which could conceivably be a causal consequence of vaccination. Thus, VAERS data must be appropriately analysed to fulfill its function, which is to generate early potential safety signals rather than to substantiate a causal relationship.

In this paper, we are using a reporting odds ratio (ROR) analysis[16] to compare the likelihood that an AEFI reported to VAERS is one of a number of neuroautoimmune AEFIs between COVID-19 and non-COVID-19 vaccines, concluding that a strong association exists in favour of COVID-19 vaccines. Based on the evidence as it presently stands, the odds of reporting a neuroautoimmune AEFI vis-a-vis any other AEFI are significantly lower than for comparable vaccines, attesting to the safety of COVID-19 vaccines.

## 2. Materials and Methods

Data was obtained for 2015 to 2021, inclusive, from the VAERS website (https://vaers.hhs.gov) on 12 June 2021. The data comprises all reports that were received between 01 January 2015 and 28 May 2021, comprising altogether 2,528,763 individual reports. Of these, 1,323,178 (52.33%) pertained to a COVID-19 vaccine.

### 2.1. Categorisation of cases

Following ingestion using Python v.3.7.5 and `pandas` v.1.2.4,[17] the resulting data frames were joined and reshaped to yield individual entries per reported symptom. Using the symptom description, reports of the following VAERS symptoms (i.e. symptoms using the coding phraseology as present in VAERS) were coded as involving a neuroautoimmune AEFI:

- Demyelinating polyneuropathy
- Immune-mediated neuropathy
- Axonal neuropathy
- Axonal and demyelinating neuropathy
- Chronic inflammatory demyelinating polyradiculoneuropathy
- Subacute inflammatory demyelinating polyradiculoneuropathy
- Autoimmune neuropathy
- Autonomic neuropathy
- Guillain-Barre syndrome
- Acute disseminated encephalomyelitis
- Demyelinating polyneuropathy
- Neuromyelitis optica spectrum disorder
- Neuromyelitis optica
- Myelitis transverse
- Multiple sclerosis relapse
- Multiple sclerosis
- Relapsing multiple sclerosis
- Progressive multiple sclerosis
- Relapsing-remitting multiple sclerosis
- Optic neuritis
- Immune-mediated encephalitis
- Autoimmune encephalopathy
- Encephalitis autoimmune
- Autoimmune demyelinating disease

Based on this encoding, the individual data was segmented and age distributions were calculated (Figure 1). Subjects receiving the COVID-19 vaccine were on average older (*µ* =50.2, *σ* = 17.8) than those who received other vaccines (*µ* =36.9, *σ* = 28.3), a phenomenon attributable to the relatively large number of childhood vaccines in relation to vaccines received in later life. Except for male recipients of the COVID-19 vaccine, age distribution of neuroautoimmune disorders followed a bimodal pattern, peaking in the third and eight decades of life. Male recipients of the COVID-19 vaccine who reported a neuroautoimmune AEFI tended towards a single mode in the early seventh decade of life.

**Figure 1.**
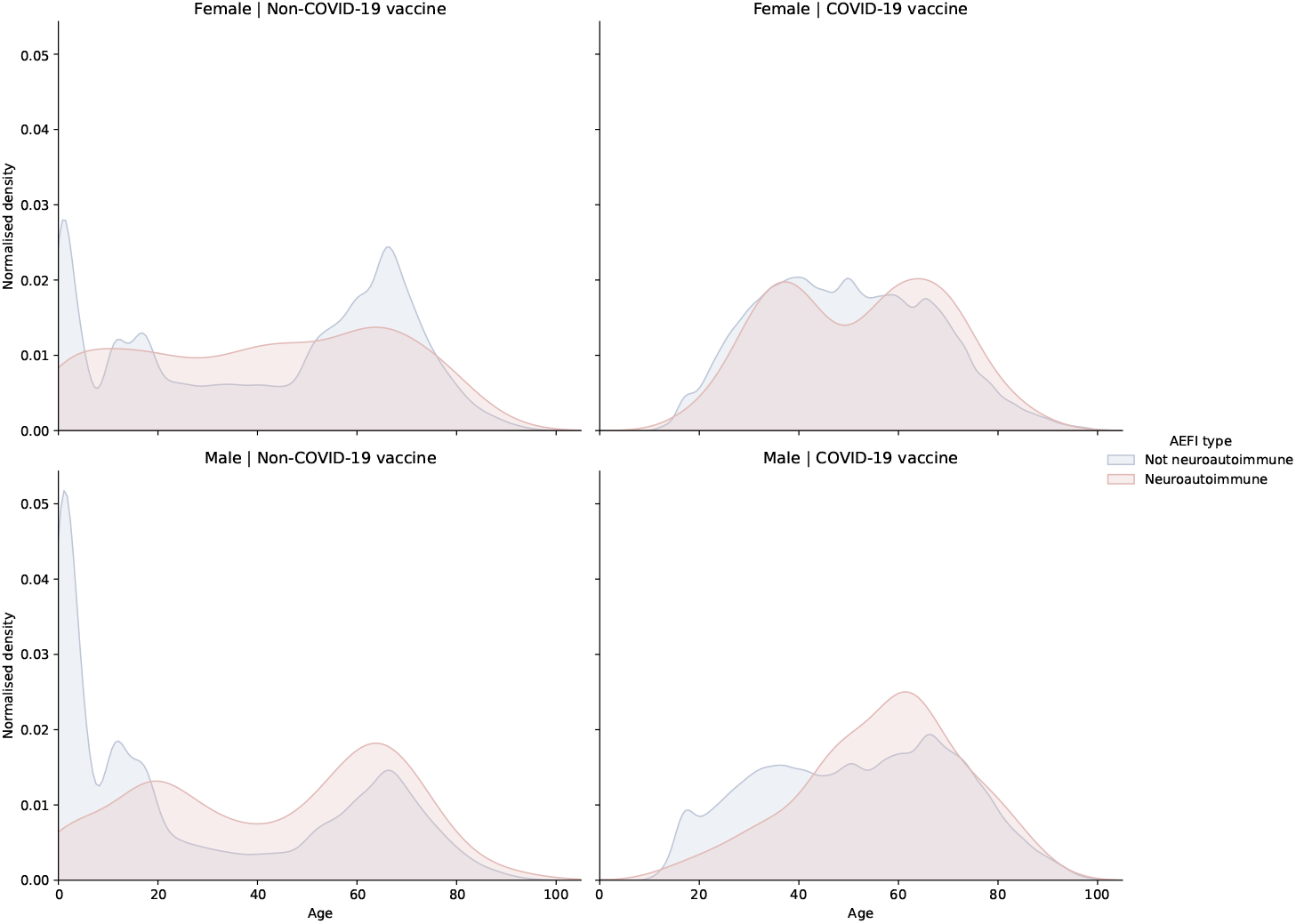
Distribution of reports by age, partitioned by vaccine type (COVID-19 vs. non-COVID-19 vaccine) and neuroautoimmune disorder status (neuroautoimmune vs. non-neuroautoimmune symptoms).

Data was cross-tabulated to yield a 2×2 contingency table for COVID-19 vs. non-COVID-19 vaccines (Table 1), and the odds ratio was calculated using Fisher’s exact test. In addition, a Pearson’s *χ*^2^ test was performed and the Pearson residuals were calculated for each permutation, using `statsmodels` v.0.12.2.[18]

**Table 1.**
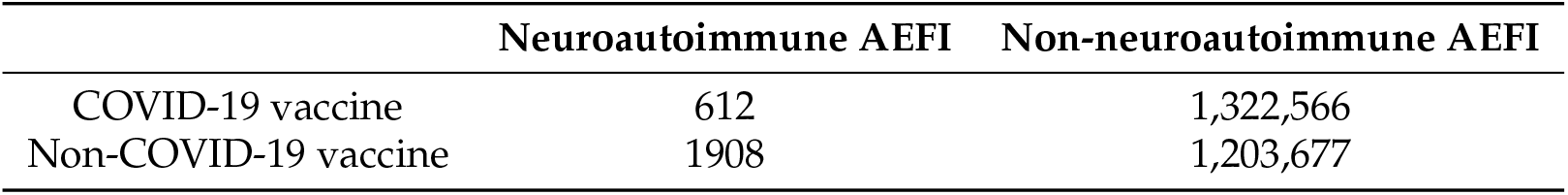
2×2 contingency table by vaccine type (COVID-19 vs. non-COVID-19 vaccine) and neuroautoimmune disorder status.

Next, symptoms were broken down individually, and a *χ*^2^ distribution fitted. Along with the actual values, the expected values under the null hypothesis of independence (*H*_0_) were ascertained, as well as the Pearson residuals.

Finally, a large contingency table was constructed, calculating the reporting odds ratio of each AEFI for each vaccine category (VAERS variable `VAX_TYPE`), which acts as a superset comprising vaccines that have typically the same target and same valency (e.g. the Johnson & Johnson COVID-19 vaccine and the Moderna vaccine are both in the COVID19 vaccine category, whereas a trivalent and a quadrivalent influenza vaccine would be different vaccine categories due to their difference in valency). For the purposes of constructing this table, a regular expression was used to filter out a number of entities that are typically recorded in VAERS but which do not indicate an appropriate denominator for the calculation of the odds ratio. These were used to exclude

- normal results (e.g. .*(`negative|normal`)$),
- mere tests and assays that do not disclose a result (e.g. .*`assay`$),
- procedures (e.g. .*(`plasty|insertion|tomy|ery…`),
- management of an ongoing medical status or device,
- administrative flags (e.g. `Blood (group|don(or|ation`))), and
- COVID-19 related public health interventions (e.g. COVID-19 (`prophylaxis|immunisation|screening`)).

RORs were cross-tabulated by vaccine type and filtered only for symptoms that were categorised as neuroautoimmmune symptomatic presentations.

## 3. Results

### 3.1. Association with neuroautoimmune AEFIs

Fisher’s two-sided exact test was used to calculate the odds ratio to determine the odds ratio for an association between reporting a neuroautoimmune AEFI and reporting after/in relationship with a COVID-19 vaccine. This yielded an OR of 0.292 at *p* < 0.0001- a statistically highly significant result that indicates COVID-19 vaccination predisposes to a significantly lower risk – by over 70% – of reporting a neuroautoimmune side effect rather than any other side effect. It is important to note that this does not mean that neuroautoimmune side effects are 70% less frequent, but rather that the likelihood of reporting a neuroautoimmune side effect over any other side effect is 70% lower.

This is confirmed by the Pearson *χ*^2^ residuals that were calculated (see Table 2). The residual of neuroautoimmune AEFIs following a COVID-19 vaccine (*−*19.459) indicates that the expected value is significantly higher than the actual value under the assumption of the null hypothesis of independence. It is therefore quite evident that the odds of reporting a neuroautoimmune AEFI over any other AEFI are correlated with whether the preceding vaccine was a COVID-19 vaccine or not, with recipients of COVID-19 vaccines being significantly (70%) less likely to report a neuroautoimmune AEFI.

**Table 2.**
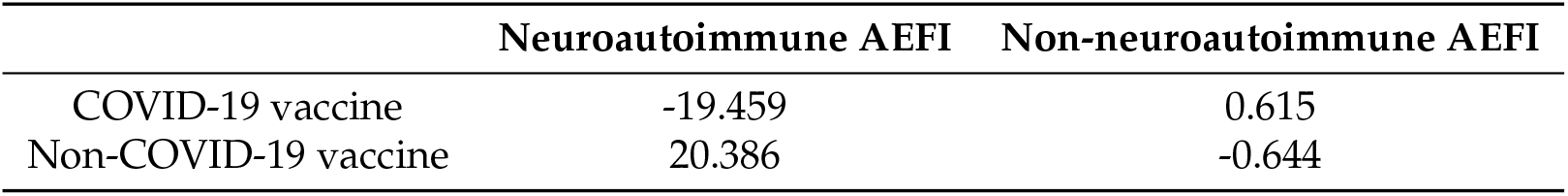
Pearson residuals by vaccine type (COVID-19 vs. non-COVID-19 vaccine) and neuroautoimmune disorder status.

|  | Neuroautoimmune AEFI | Non-neuroautoimmune AEFI |
| --- | --- | --- |
| COVID-19 vaccine | -19.459 | 0.615 |
| Non-COVID-19 vaccine | 20.386 | -0.644 |

### 3.2. Association of individual neuroautoimmune AEFIs

Following on from these findings, it is not surprising to see that in most cases, the number of actual cases reported is significantly lower than would be expected under the null hypothesis of independence (see Figure 2). The notable exceptions are

**Figure 2.**
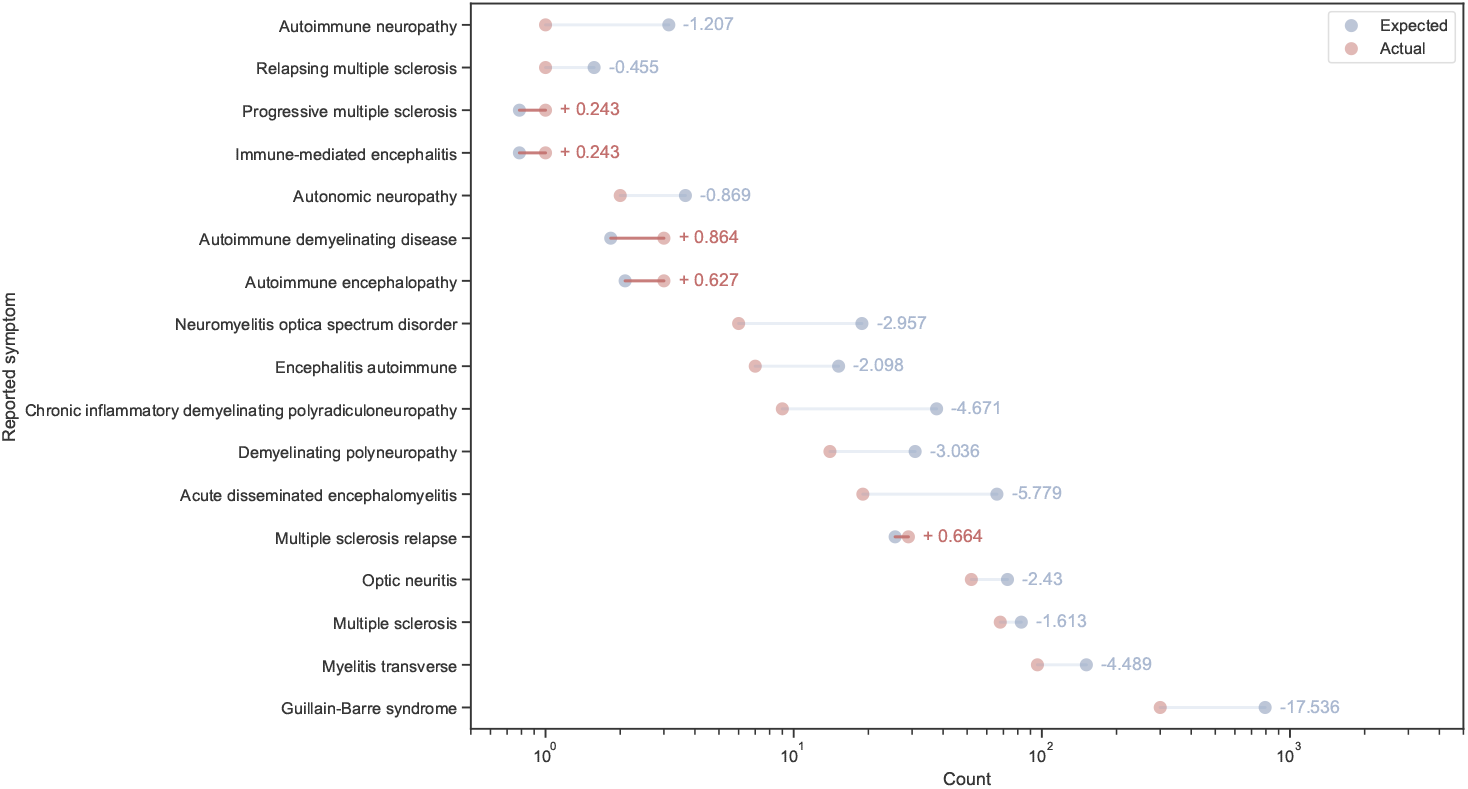
Expected vs. actual number of reports for individual neuroautoimmune symptoms based on Pearson’s *χ*^2^ test along with Pearson residuals for COVID-19 vaccines.

- progressive MS (Pearson residual: 0.243),
- immune-mediated encephalitis (Pearson residual: 0.243),
- autoimmune demyelinating disease (Pearson residual: 0.864),
- autoimmune encephalopathy (Pearson residual: 0.627), and
- MS relapse (Pearson residual: 0.664).

While the residuals indicate that the deviation is quite small for all of the above, it appears that in contrast with other vaccines, COVID-19 vaccines appear to result in a relatively higher number than expected of these AEFIs. This must be considered in the context of actual figures, of course – altogether, these differences account for fewer than 0.25 additional cases of progressive MS and immune-mediated encephalitis, fewer than one case of autoimmune encephalopathy, fewer than 4 cases of MS relapse and fewer than 1.5 cases of autoimmune demyelinating disease. Of over 1.3m reports, these constitute an excess of six cases, or approx. one in 220,530 reports. Consequently, it remains to be seen (especially given the low absolute numbers) whether this association remains in evidence as more data accrues, or, as is more likely, it is part of the noisy undercurrent that is characteristic of passive reporting.

### 3.3. Reporting odds ratios between vaccines

As Figure 3 shows, the ROR of neuroautoimmune presentations is significantly lower for COVID-19 vaccines than it is for any of the comparator vaccines. The group of comparators was chosen to primarily include vaccines that have the same age range of approved users to reduce the bias arising from the fact that some vaccines are typically given in a paediatric context (e.g. MMR), while COVID-19 vaccination was consciously designed to address older populations first due to their higher overall risk from COVID-19 as well as the relatively higher rate of respiratory, cardiac and endocrine comorbidities that governed vaccine allocation in the beginning.

**Figure 3.**
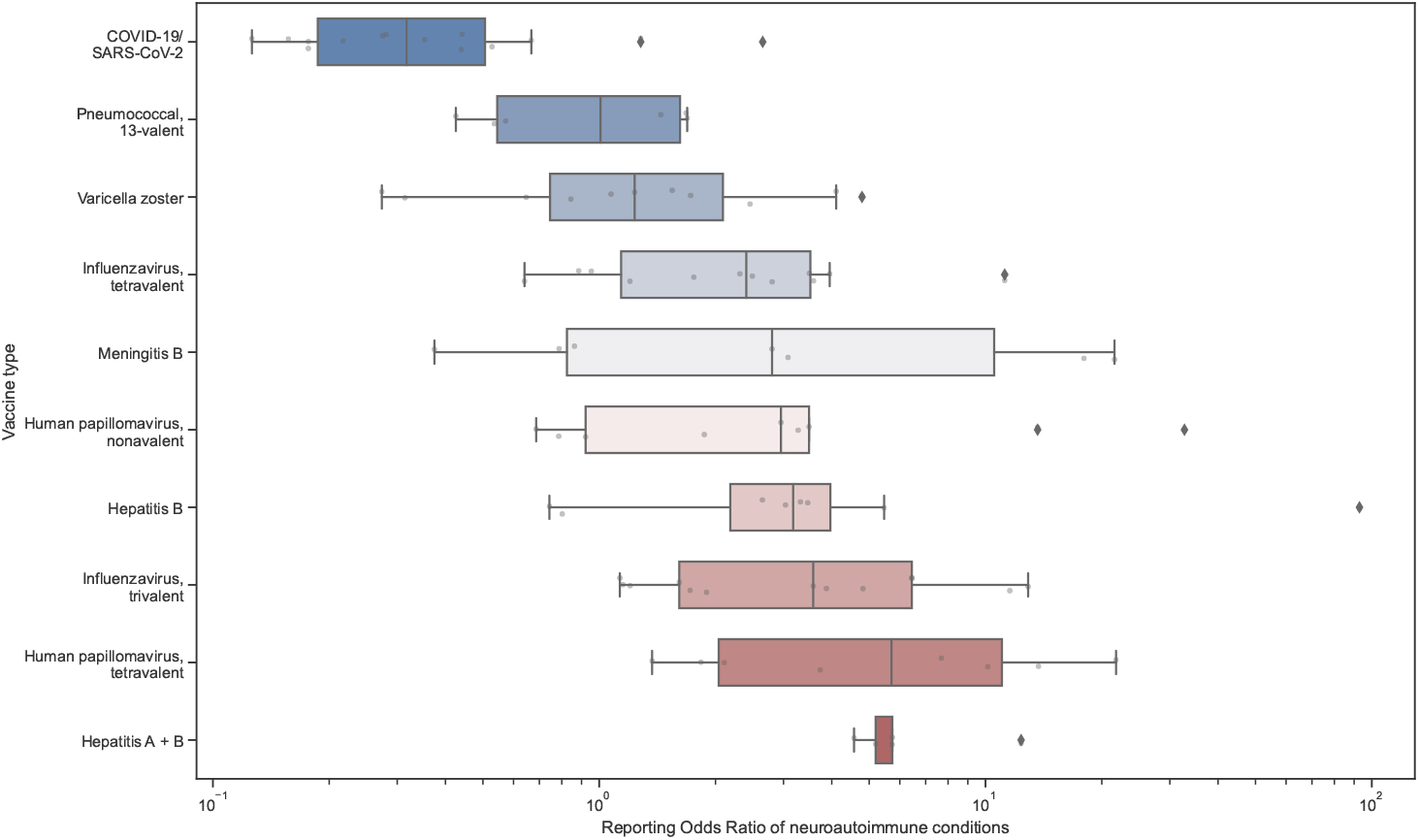
Reporting Odds Ratios of neuroautoimmune conditions in a range of vaccines approved for adult populations.

## 4. Discussion

Based on early data from VAERS for reports about COVID-19 vaccinations received on or before 28 May 2021, there is convincing evidence that COVID-19 vaccines compare remarkably well with other vaccines in respect of neuroautoimmune AEFIs. Despite the limitations of this study, which drew on passive reporting data with all its inherent problems, such as reporting bias and the lack of a reliable denominator, we have found evidence that supports the safety of COVID-19 vaccines with regard to neuroimmunological presentations. Statistical analysis indicated that in comparison to recipients of other vaccines, those who reported in respect of a COVID-19 vaccines were over 70% less likely to report a neuroimmunological presentation than a non-neuroimmunological presentation.

Similarly, the number of reports fell significantly under what was expected in all but two somemwhat cohesive symptom groups: relapses of MS on one hand and autoimmmune encephalitis on the other. The fact that some forms of autoimmune encephalitis are distributed bimodally by age with a peak in the third and the seventh decade of life may support a hypothesis that the increase in cases of autoimmune encephalitis is the consequence of statistical noise and/or sampling bias due to the higher average age of COVID-19 vaccine recipients in the sample.[19] Similarly, multiple sclerosis exhibits a similar bimodality of age distribution, to the point that respective terms for early-onset (teens to early 30s) and late onset (50s and later) MS have become established in the literature.[20] Nevertheless, clinicians may, as a matter of abundance of caution, keep a closer eye on patients with known MS or risk factors for MS (such as previous diagnosis of optic neuritis or CIS/RIS) for signs of progression or relapse. This may be accompanied by proactively explaining the relative risk of a relapse in the context of potentially worse clinical outcomes with COVID-19, especially for patients treated with systemic immunosupressants.

Since the start of the global vaccination campaign, evidence has been accumulating for the overall tolerability and safety of the COVID-19 vaccines. This study seeks to add to our current understanding by quantifying the highly favourable risk profile of COVID-19 vaccines with regard to neuroautoimmune disorders. While some questions remain open and conclusions made over early data may have to be reconfirmed through ongoing analysis, the current data supports the assertion that the COVID-19 vaccine has perhaps the lowest reporting odds ratio for neuroautoimmune disorders of any vaccine routinely given in adulthood. This is a reassuring indication of safety at a time when concerns about adverse events still keep most countries from attaining the threshold of collective immunity.

## Data Availability

Source data can be obtained from the FDA/CDC's VAERS site. All code is publicly available on Github and under the DOI 10.5281/zenodo.4940261.

https://vaers.hhs.gov/data.html

https://github.com/chrisvoncsefalvay/covid19-neuroautoimmune-aefis

## Funding

This research was funded by Starschema Inc. under its intramural research funding programme.

## Data Availability Statement

VAERS reporting data is available from the CDC’s website at https://vaers.hhs.gov. All code and scripts supporting this manuscript are deposited at https://github.com/chrisvoncsefalvay/covid19-neuroautoimmune-aefis and are made available under the DOI 10.5281/zenodo.4940261.

## Conflicts of Interest

CvC is a consultant to a company that may be affected by the research reported in this paper. The funders had no role in the design of the study; in the collection, analyses, or interpretation of data; in the writing of the manuscript, or in the decision to publish the results.

### Abbreviations

The following abbreviations are used in this manuscript:

AEFI: Adverse event following immmunization
CDC: Centers for Disease Control and Prevention
CIS: clinically isolated syndrome
EUA: Emergency Use Authorization
FDA: Food and Drug Administration
GBS: Guillain-Barre syndrome
mRNA: messenger RNA
MS: multiple sclerosis
QoL: quality of life
RIS: radiologically isolated syndrome
ROR: Reporting Odds Ratio
VAERS: Vaccine Adverse Effect Reporting System

## Notes

### Author Declarations

No IRB approval has been sought as the study involves publicly available data released by the CDC/FDA.

### Summary of Updates

Contingency tables were reformatted to present the contingency case counts and Pearson's residuals in a more readable manner.

